# Anxiety and depression among the adult population with prior infections of dengue with and without COVID-19

**DOI:** 10.1101/2025.06.13.25329554

**Authors:** Raju Ghimire, Mamata Pradhan, Keshab Parajuli, Tara Ramel

**Author notes:** **Corresponding author** “RG”. RG is the senior author, and all other authors contributed equally to this work.

## Abstract

Several separate studies of impact of dengue and COVID-19 on mental health revealed that both the infections possesses severe impact on the mental health and wellbeing of people. There are very few studies showing the mental health status with the people having dengue infection and the mental health status with the people having dengue with COVID-19. The study aims to find out the mental health status of the people having dengue infection with and without COVID-19 infection.

The study was community based Comparative cross-sectional study design, in the adult population of age 18 and above in one municipality selected randomly of each district at Kathmandu valley. The sample size of the study is 598 calculated by using cross-sectional comparative two-independent sample study formula.

DASS 21 tool was administered to eligible participants through face-to-face interview. Monovariate, Bivariate (Chi-square test, and multivariate analysis (bivariate logistic regression) was done.

In bivariate analysis age (p=0.003), ethnicity (0.001), education level (p= 0.005), have chronic disease (p=0.016) showed significant association of anxiety due to both dengue with covid 19 infection. Similarly religion (p= 0.0044), Ethnicity (p= 0.001), Physical exercise (p= 0.015), habit of consumption of fruit (p=0.033) showed significant association with occurance of depression due to prior infection of dengue with covid 19. In multivariate analysis (binomial regression analysis) no significant association of infection of dengue with covid 19 was found with any independent variables while anxiety due to infection of dengue with covid 19 was found to be significant with ethnicity 0.004( AOR=2.538), habit of smoking of smoking 0.049 was found.

The study demonstrated that prior infections of dengue, with and without COVID-19, significantly influence mental health outcomes in adults in the Kathmandu Valley. Mental health services in health care setting is recommended at policy level and implemented at local level.

## Introduction

Dengue is a mosquito-borne viral disease caused by dengue virus that has rapidly spread to all regions of WHO in recent years. (1) Dengue fever is also recognized as break bone fever. (2) Dengue is found in tropical and sub-tropical climates worldwide, mostly in urban and semi-urban areas. (3) Dengue fever is one of the major infectious diseases and a serious public sector health concern. (4) The COVID-19 pandemic has had a severe impact on the mental health and wellbeing of people around the world. (5) According to a scientific brief released by the World Health Organization (WHO), during the first year of the COVID-19 pandemic, global prevalence of anxiety and depression increased by a massive 25%, (6)

WHO report shows global incidence of dengue has grown dramatically with about half of the world’s population now at risk. Although an estimated 100-400, million infections occur each year. The majority (60%) of the world’s population is predicted to be at risk by 2080.It is endemic in 129 countries with 70% of cases in Asia. (3) Because of rapid increase in incidence and widespread prevalence, dengue has become a major public health concern. The WHO launched “The Global Strategy for Dengue Prevention and Control 2012–2020” to address the threat of an impending pandemic and reduce the burden of dengue. (7)

Evidence shows physical condition of individuals greatly affects their psychological health. Patients suffering from different diseases also start suffering from emotional problems like depression, anxiety, stress and other psychiatric conditions. (4) Neurological and especially psychiatric manifestations are increasingly reported in acute as well as convalescent phases of dengue infection. (8) Patients suffering from dengue fever also develop psychiatric and serious psychological conditions like phobias and post-traumatic stress disorder. They see huge death toll around them and develop a fear of death. (4) Despite the high prevalence of this illness, the exact incidence of neuropsychiatric manifestation of Dengue infection is not certain due to the lack of adequate studies. (9)

According to the World Health Organization, Nepal reported its first dengue case in a traveler returning from India in 2004. Since then, dengue has been endemic in Nepal. Nepal is facing outbreak of dengue this year. Cases have been reported from all seven provinces, affecting all 77 districts in the country. Dengue cases have increased since July coinciding with the rainy season with the majority of the cases reported during September (83.6 percent). The highest number of new cases in 2022 has been reported in the districts of Kathmandu (33.8 percent), Lalitpur (23.2 percent), and Makawanpur (9.8 percent). (3)

The 2022 outbreak of dengue shows post-acute dengue syndrome is becoming apparent and drawing that required more attention in Nepal. Post-acute dengue syndrome has not yet been widely recognized in Nepal. In light of the current dengue outbreak in Nepal, the sudden rise in post-acute dengue syndrome can be considered as an unexpected but possibly a new emerging event. (11)

The study done in several fever clinics across Nepal on May—June, 2020 found that increased prevalence of stress, anxiety and depressive symptoms during the initial stage of COVID-19 pandemic in Nepal. The study shows prevalence of anxiety; depression and stress were 14%, 7% and 5% respectively. (12)

The COVID-19 pandemic may have brought many changes to how people live their life. They have uncertainty, altered daily routines, financial pressures and social isolation. People may worry about getting sick, how long the pandemic will last, whether their job will be affected and what the future will bring. Information overload, rumors and misinformation can make people life feel out of control and make it unclear what to do. (10)

Anxiety and depression which are often omitted by patients and their family are overlooked and neglected by the physicians as well. Psychiatric assessment of patients suffering from DF and initiation of appropriate treatment will facilitate psychosocial adaptation of the patients and early recovery. (2) As Dengue is endemic in Nepal, there is urgent need to conduct studies in order to assess the impact of Dengue on mental health and important to evaluate the burden of the disease on mental functioning. The untreated depression can lead to increased medical morbidity, higher healthcare costs, and can delay or prevent recovery from medical illness. This study aimed at assessing the level of anxiety and depression among the adult population with prior infections of dengue with and without COVID-19 in Kathmandu valley and the study found heightened anxiety and depression who were infected with both dengue and COVID-19 than those who were infected with dengue only.

## Materials and Methods

### a. Research design

A cross-sectional comparative study design was used in this research.

### b. Description of research design

This study was descriptive quantitative cross-sectional study. A quantitative approach was applied to measure the association between the level of anxiety and depression among the adult population with prior infections of dengue with and without COVID-19.

### c. Study area

The study was conducted in Kathmandu valley i.e. Lalitpur, Bhaktapur and Kathmandu.

### d. Study site and Justification

The study was conducted in at least one metropolitan city/municipality from each district within Kathmandu valley with a high number of dengue and COVID-19 cases.

### e. Study population

The study population was adults of age above 18 years who were residents of Kathmandu valley (at least six months) and were confirmed with dengue infection (sample 588). For the comparison purpose, out of all selected dengue confirmed cases 50% of the people (294) were previously infected with COVID-19 infected within 3 years i.e. between Jan 2020-Dec 2022.

**Study team:** Three public health researchers and one mental health expert were involved in the study.

### f. Sampling Unit

Adult population above 18 years who are residents of Kathmandu valley.

### g. Sample Size

To calculate the sample size, cross-sectional comparative two-independent sample study formula was used. The following parameters were used to calculate the sample size;

Incidence, group 1= 50% assumed (having dengue who were previously infected with covid-19)

Incidence group 2= 25% assumed (having dengue who were not previously infected with covid-19)

Alpha = 0.05

Beta= 0.2

Power= 80%

Sample size

Group 1-267

Group 2-267

Total sample= 267+267= 534

After adding 10% nonresponse rate to the calculated sample size, the final sample size is 588.

### h. Sampling Technique

Kathmandu valley is selected purposefully. Simple random sampling was employed to select the municipalities. The wards were selected using simple random sampling. An equal sample was taken from each ward. (As we do not have concrete ward-wise data on the number of people infected with COVID and dengue, we propose to include an equal number of participants from each ward). To select the household in the ward, the researcher (data collector) went to that ward’s centre/popular marketplace; then, spined the pen or bottle and follow the direction of the tip. The data collector visited the first household and asked the adult population about their disease history. The neighbour comparison was selected for the study. The next adjoining house of the participants with prior dengue infections without COVID-19 was chosen for the comparison. Only a participant from a house was taken for the study. When the data collector reached the end of the ward, he/she turned the right side for the additional samples from each ward.

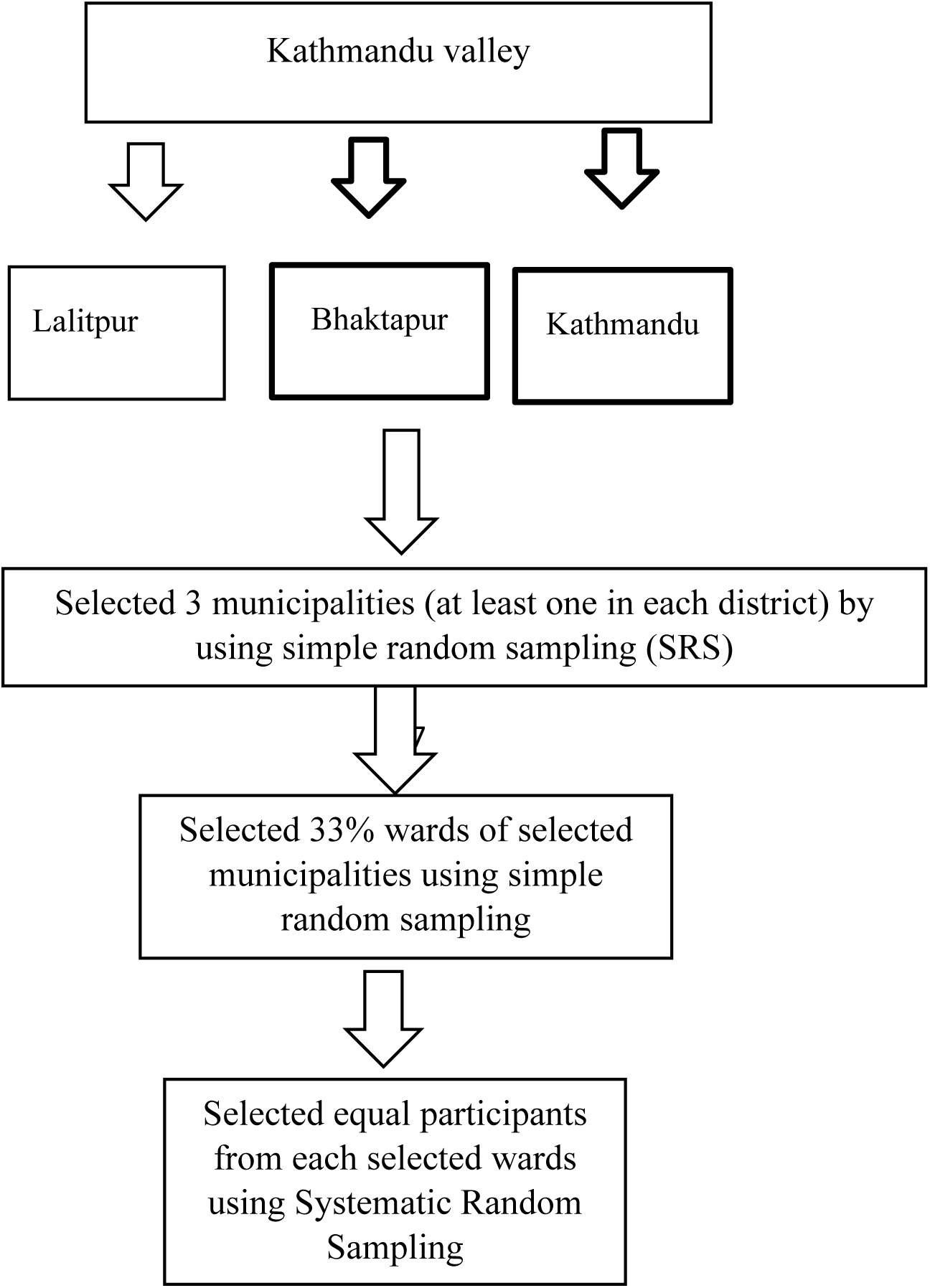

### i. Criteria for sample selection

Adult population of selected ward were selected for research participants. Sample was selected through systematic random sampling technique.

### j. Data collection techniques

The purpose of the research was explained to each respondent. Data was collected by using a structured interview schedule through face-to-face interview method.

### k. Data collection tools

A structured questionnaire was developed to collect the data on the socio-demographic characteristics of the sampled population.

The validated translated tool of DASS-21 was used to measure anxiety and depression in the study population. Both the anxiety and depression scale contain 7 questions each.

The Depression, Anxiety and Stress Scale - 21 Items (DASS-21)(16) is a set of three self-report scales designed to measure the emotional states of depression, anxiety and Depression, Anxiety and Stress Scale - 21 Items (DASS-21). Individuals required to rate the presence of each symptom during the infection period. Each subscale consisted of seven items, and each item is rated on a Likert type scale ranging from 0 “did not apply to me at all”–3 “applied to me all the time.”

### l. Data Analysis

Data was entered in Epi-data version 3.1 where entry, editing and checking of data was done. Descriptive analysis (mean, frequency and percentage) was carried out. First of all, the demographic characteristics of the sample was assessed. Then, bivariate chi-square analyses were examined to find out the associations between demographic. For the comparison of outcome among the covid infected and non-infected adults having dengue, non-parametric

Mann Whitney U test was used. After that the estimated prevalence of depression and anxiety was measured, as defined by the standard tool cutoff scores on each measure. Lastly, after controlling for demographic characteristics (i.e., age, race/ethnicity, and gender and co-morbidities), multivariate logistic regression was calculated to estimate odds ratios at 95% confidence intervals for the dengue-related stressors and significant levels of depression and anxiety. All statistical analyses were carried out by using SPSS.

### m. Exclusion and Inclusion Criteria

Inclusion criteria: -

All the participants who were residing in the study area regardless of permanency (those residing in the area for more than 6 months) were included in the study.

Adult population above 18 years were included in the study.

**Exclusion:**-

Those who didn’t provide consent for data collection from among the inclusion population were excluded from the study.

### n. Validity and Reliability of tools

To ensure the reliability and validity, questionnaire was constructed by consulting respected guide. Pretesting of tools was done to 10% of population before data collection. Cross checking was done in order to reduce the error.

### o. Potential Biases

- Recall bias
- Interview bias

In order to minimize the recall bias standard tool was used and the participants were made to understand the question by telling the question for more than 1 time and tried to correlate the issues with the things they know or remember.

In order to reduce the interview bias interviewer

- Read each question exactly as it appeared.
- Didn’t interpret the question for the interviewee.
- Offer to repeat the question exactly as it appeared

### p. Ethical Issues

Ethical approval was taken from the Nepal Health Research Council (NHRC). Written permission from each district public/ health office was taken. Before the interview, written informed consent was taken from the participants for their participation in the research and offered justifications. Participants voluntarily participated in answering the questions and were provided the right to deny any time during data collection. The respondents were assured that the answer given remained private, unanimous and confidential and only used for study purpose.

## Result

### Description of Socio-demographic Information

**Table 1.** Information on infection of dengue and covid-19.

| Variable | Frequency | Percent (%) |
| --- | --- | --- |
| **Suffer from dengue disease between 3 years** |  |  |
| Yes | 598 | 100 |
| **Suffer from both dengue and covid-19 within 3 years** |  |  |
| Yes | 296 | 49.5 |
| No | 302 | 50.5 |

The table shows that almost all 598 (100%) have had dengue disease. Among the 598 respondents with dengue; nearly half of the respondents 296 (49.5%) have suffered from both dengue and COVID-19, while 303 (50.5%) had not had covid 19 infection in the past three years.

Total number of participants was 598. Participants were from minimum of 18 years to maximum of 80 years with mean age of 39.90 years, median age 35 years and S.D of 15.66 years.

**Table 2.** Socio-demographic Information.

| Variable | Frequency | Percent (%) |
| --- | --- | --- |
| **Gender** |  |  |
| Male | 278 | 46.5 |
| Female | 320 | 53.5 |
| **Religion** |  |  |
| Hindu | 514 | 86 |
| Christian | 24 | 4 |
| Buddhist | 52 | 8.7 |
| Others | 8 | 1.3 |

The table indicates that out of 598 respondents, more than half 320 (53.5%) were female, while the remaining 278 (46.5%) were male. Among the respondents, the majority, 516 (86%), are Hindu. The remaining respondents are Christian (24 or 4%), Buddhist (52 or 8.7%), and other religions (8 or 1.3%).

**Table 3.** Socio-demographic Information.

| Variable | Frequency | Percent (%) |
| --- | --- | --- |
| **Ethnicity** |  |  |
| Dalit | 21 | 3.5 |
| Under Privileged | 38 | 6.3 |
| Under Privileged Madhesi | 11 | 1.8 |
| Privileged minority | 13 | 2.2 |
| Privileged janajati | 388 | 65 |
| Bhramin/Chhetri | 127 | 21.2 |

The table indicates that out of 598 respondents, 388 (65%) were Privileged Janajati, 127 (21.2%) were Brahmin/Chhetri, and the remaining were Dalits 21(3.5%), Under Privileged 38(6.3%), Under Privileged Madhesi 11(1.8%), and Privileged Minority 13(2.2%).

**Table 4.** Socio-demographic Information.

| Variable | Frequency | Percent (%) |
| --- | --- | --- |
| **Marital Status** |  |  |
| married | 358 | 59.8 |
| Unmarried | 194 | 32.5 |
| Widow | 39 | 6.5 |
| Divorced | 3 | 0.5 |
| Living Separate | 4 | 0.7 |
| **Educational Level** |  |  |
| Illiterate | 71 | 11.8 |
| Primary 1-8 | 81 | 13.5 |
| Secondary 8-10 | 74 | 12.5 |
| High secondary 11-12 | 190 | 31.8 |
| Bachelor degree | 162 | 27 |
| Above Bachelor | 20 | 3.3 |

The table indicates that out of 598 respondents, more than half (358 or 59.8%) are married, one third (194 or 32.5%) are unmarried, 39 (6.5%) are widows, and 3 (0.5%) and 4 (0.7%) are divorced and living separately, respectively. Regarding education, one third (191 or 31.8%) have a high secondary education (grades 11-12), 162 (27%) have a bachelor’s degree, 71 (11.8%) are illiterate, 81 (13.5%) have a primary education (grades 1-8), 75 (12.5%) have a secondary education (grades 8-10), and 20 (3.3%) have education above a bachelor’s degree.

**Table 5.** Socio-demographic Information.

| Variable | Frequency | Percent (%) |
| --- | --- | --- |
| **Household monthly income** |  |  |
| >10,000 | 13 | 2.2 |
| 11,000-20,000 | 40 | 6.7 |
| 20,000-30,000 | 124 | 20.7 |
| 30,000-40,000 | 182 | 30.3 |
| 40,000-50,000 | 144 | 24.2 |
| 50,000> | 95 | 16 |

| **Employment status** |  |  |
| --- | --- | --- |
| Yes | 398 | 66.5 |
| No | 200 | 33.5 |
| **Household saving** |  |  |
| Yes | 468 | 78.3 |
| No | 130 | 21.7 |

The table shows that one third 182 (30.3%) of respondents have a household monthly income of 30,000-40,000, while 144 (24.2%) have an income of 40,000-50,000, 124 (20.7%) have 20,000-30,000, 95 (16%) have more than 50,000, 40 (6.7%) have 11,000-20,000, and the remaining 13 (2.2%) have less than 10,000. Additionally, one third 201(33.5%) have no employment status, while 399 (66.5%) are employed. Among all respondents, the majority 468(78.3%) have household savings, while the remaining 130 (21.7%) do not have any household savings.

**Table 6.** Health habits related questions.

| **Variable** | **Frequency** | **Percent (%)** |
| --- | --- | --- |
| **Have habit of smoking** |  |  |
| Never | 430 | 71.8 |
| Occasionally | 115 | 19.3 |
| Regular | 53 | 8.8 |
| **Have habit of alcohol** |  |  |
| Never | 295 | 49.5 |
| Occasionally | 295 | 49.2 |
| Regular | 8 | 1.3 |

The table indicates that out of 598 respondents, 430 (71.8%) have never smoked, 115 (19.3%) smoke occasionally, and 53 (8.8%) are regular smokers. Additionally, around half 295 (49.5%) have never consumed alcohol, 295 (49.2%) drink occasionally, and 8 (1.3%) are regular alcohol consumers.

**Table 7.** Health habits related questions.

| Variable | Frequency | Percent (%) |
| --- | --- | --- |
| **Have habit of physical exercise** |  |  |
| Yes | 277 | 46.3 |
| No | 321 | 53.7 |
| **Have habit of consumption of fruits** |  |  |
| Yes | 551 | 92.1 |
| No | 47 | 7.9 |
| **Have habit of consumption of vegetables** |  |  |
| Yes | 570 | 95.2 |
| No | 28 | 4.8 |

The table indicates that out of 598 respondents, more than half 321 (53.7%) do not engage in physical exercise, while 277 (46.3%) do physical exercise. Additionally, the maximum respondent 551 (92.1%) regularly consume fruits, whereas 47 (7.9%) do not. all most all respondents 570 (95.2%) have a habit of consuming vegetables, while 28 (4.8%) do not habit of consuming vegetables.

**Table 8.**
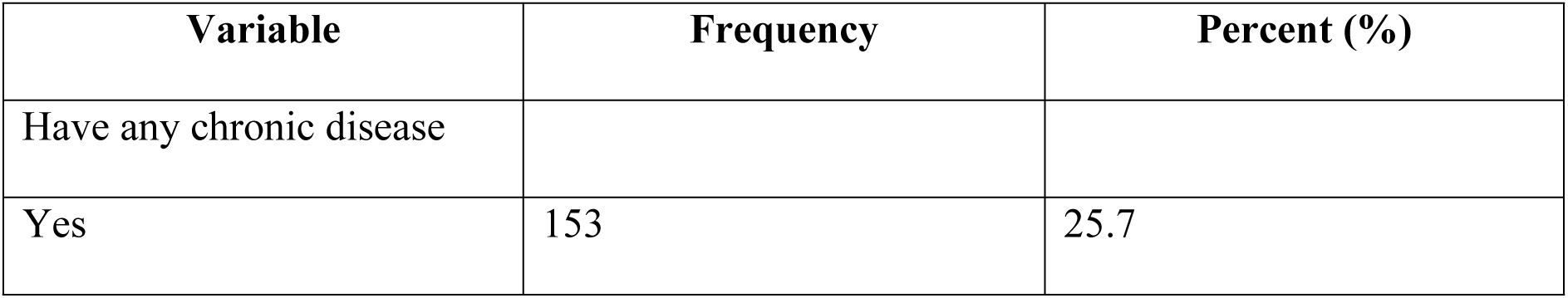
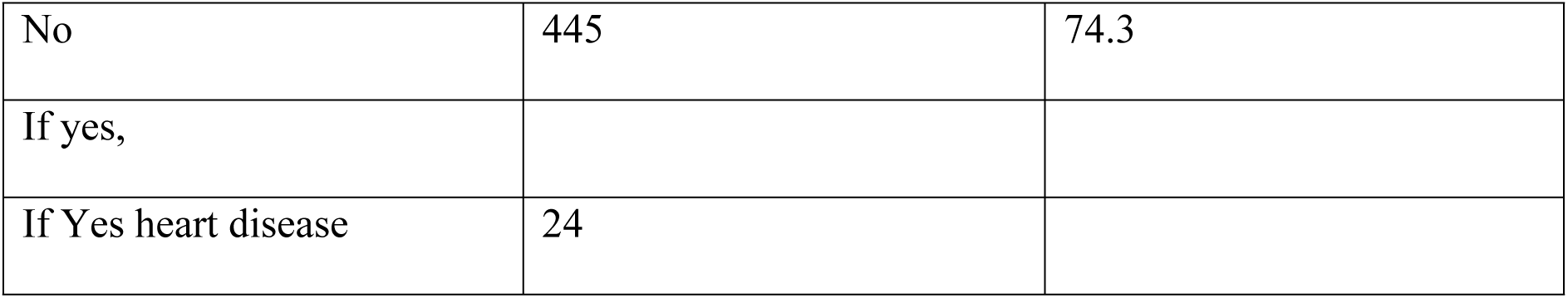
Have any chronic disease.

**Table 9.** Participation of family members.

| Variable | Frequency | Percent (%) |
| --- | --- | --- |
| **Family member time spend with you** |  |  |
| Always | 472 | 78.7 |
| Sometimes | 126 | 21 |
| Not at all | 2 | 0.3 |
| **family members involve you in decision making** |  |  |
| Yes | 550 | 91.7 |
| No | 50 | 8.3 |
| **Receive pension** |  |  |
| Yes | 54 | 9 |
| No | 546 | 91 |

The table indicates that out of 600 respondents, the majority 472(78.7%) always have family members spending time with them, 126 (21%) sometimes have family members around, and 2 (0.3%) do not have family members spending time with them at all. Additionally, maximum respondents 550(91.7%) have family members involved them in decision-making, while 50 (8.3%) do not. Furthermore, the maximum respondent 546 (91%) do not receive a pension, whereas the remaining 54 (9%) do receive a pension.

### Level of Anxiety due to Dengue

Before interpreting the scores, the summed numbers in anxiety scale is multiplied by 2 (this is because the DASS 21 is the short form of the scale) and is categorized according to the criteria defined by it.

**Table 10.** Level of Anxiety due to Dengue n=598.

| Anxiety | Frequency | Per cent (%) |
| --- | --- | --- |
| Normal | 173 | 28.9 |
| Mild | 36 | 6 |
| Moderate | 154 | 25.8 |
| Severe | 82 | 13.7 |
| Extremely severe | 153 | 25.6 |

The above table shows that there was normal, moderate and extremely severe anxiety in one fourth of the participants respectively in each category.

Before interpreting the scores, the summed numbers in depression scale is multiplied by 2 (this is because the DASS 21 is the short form of the scale) and is categorized according to the criteria defined by it.

**Table 11.** Level of Depression due to Dengue n= 598.

| Depression | Frequency | Per cent (%) |
| --- | --- | --- |
| Normal | 244 | 40.8 |
| Mild | 87 | 14.5 |
| Moderate | 131 | 21.9 |
| Severe | 89 | 14.9 |
| Extremely severe | 47 | 7.9 |

The table shows that there is normal depression in two fifth of the individual and more than two fifth of the participants were suffering from severe and extremely severe depression.

**Table 12.**
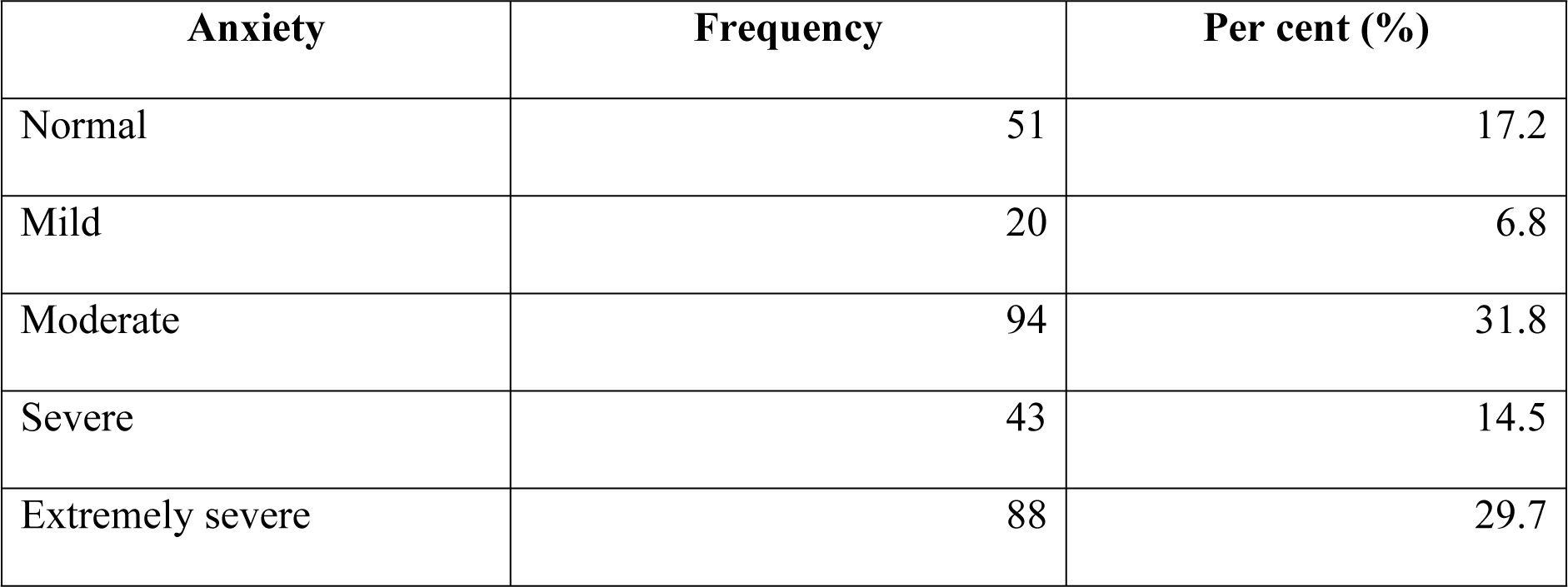
Level of anxiety among the participants who were suffered from both dengue and covid n= 296.

| **Anxiety** | **Frequency** | **Per cent (%)** |
| --- | --- | --- |
| Normal | 51 | 17.2 |
| Mild | 20 | 6.8 |
| Moderate | 94 | 31.8 |
| Severe | 43 | 14.5 |
| Extremely severe | 88 | 29.7 |

The above table showed that nearly half of the respondents (44.2%) who were suffered from both dengue and COVID had sever (severe and extremely severe) anxiety.

**Table 13.**
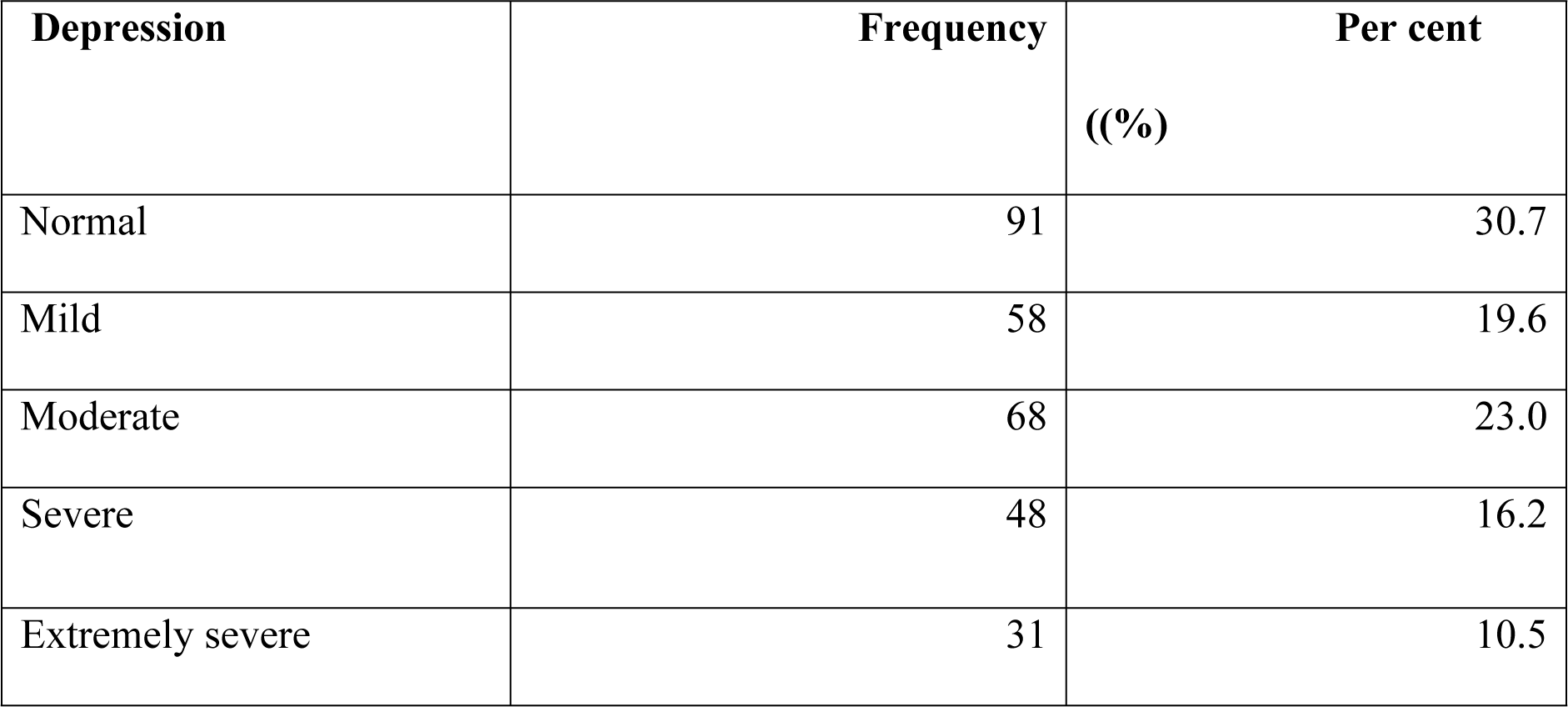
Level of depression among the participants who were suffered from both dengue and covid 19 N=296.

The result from the table showed that more than one forth (26.7%) of the respondents who were suffered from both dengue and COVID had severe (sever and extremely severe) depression.

**Figure no. 3: Comparison of level of anxiety due to dengue infection and anxiety due to both dengue and COVID 19 in percentage**

The above graph shows the comparative result of anxiety due to dengue infection and anxiety due to infection of both dengue and COVID 19.

There are 6% more moderate cases and more than 3% extremely severe cases due to infection of both dengue and COVID 19 than those with dengue infection only.

**Figure 4: Comparison of level of depression due to dengue infection and depression due to both dengue and COVID 19 in percentage**

The above figure showed the level of depression due to dengue infection only and depression due to the infection of both dengue and COIVD 19.

The above figure depicts that level of depression due to infection of both dengue and COVID 19 is slightly high in each category i.e. Mild, Moderate, Severe and extremely severe respectively than with the infection of dengue only.

### Bivariate Analysis

**Table 14.**
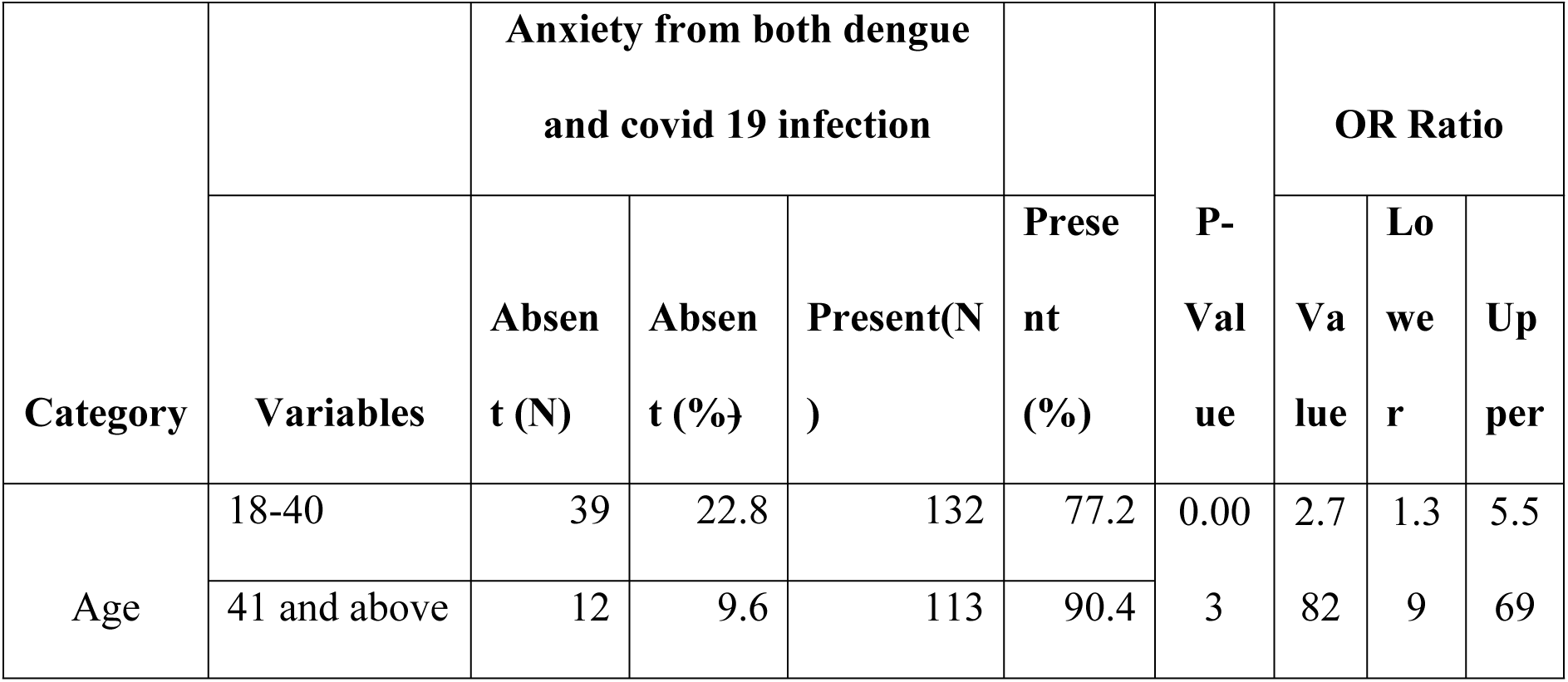
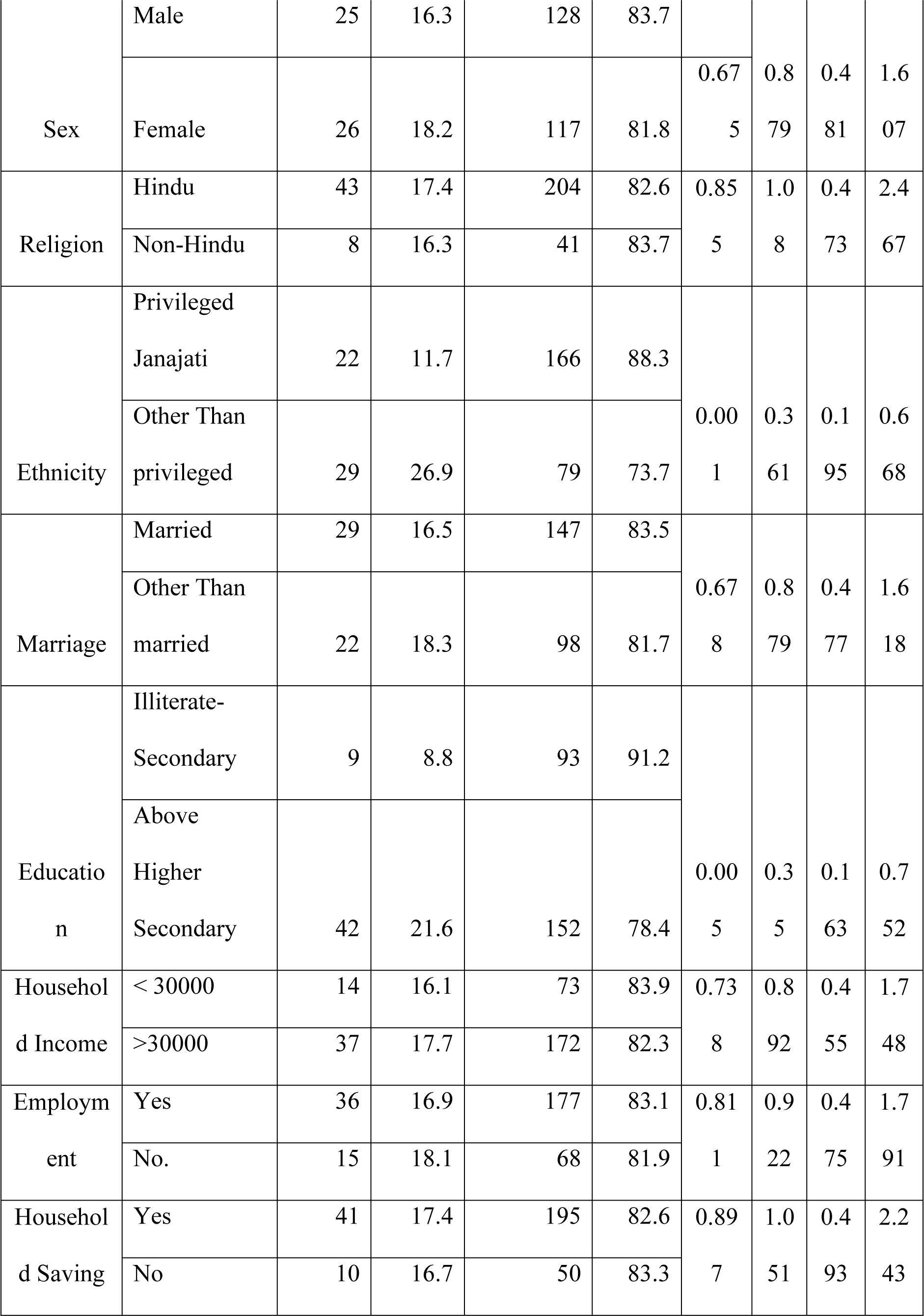
Bivariate Analysis between Socio-demographic variable and Anxiety from both Dengue and Covid-19 infection.

Above table shows significant as well as non-significant associations of anxiety with demographic and socioeconomic factors. Anxiety was significantly related to age with people 41 or older way more likely to be anxious (90.4%) than people 18–40 (77.2%) with an odds ratio of 2.782 (95% CI: 1.39–5.569, p = 0.003). Ethnicity also showed a difference in anxiety with privileged Janajati being less anxious (88.3%) than all other (73.7%), with an odds ratio of 0.361(95% CI: 0.195–0.668, p = 0.001). Education was also an independent predictive variable, as higher education was associated with significantly less anxiety (78.4 vs 91.2%) when compared to those with lower education (odds ratio: 0.35, 95% CI: 0.163–0.752, p = 0.005). Other factors, such as sex, religion, marital status, household income, job status, and household savings, were not significantly related to anxiety, as their p-values were greater than 0.05. These results indicate age, ethnicity, and education as significant by anxiety indicators in the population sampled

**Table 15.** Association between anxiety form Both dengue and covid 19 infection with health-related factor.

| Category | Variables | Anxiety form Both dengue and covid 19 infection |  |  |  | P-Value | OR Ratio |  |  |
| --- | --- | --- | --- | --- | --- | --- | --- | --- | --- |
|  |  | Absent (N) | Absent (%) | Present(N) | Present (%) |  | Value | Lower | Upper |
| Smoking Habit | Never | 39 | 19.5 | 161 | 80.5 | 0.307 |  |  |  |
|  | Occasional | 9 | 13.4 | 58 | 86.6 |  |  |  |  |
|  | Regular | 3 | 10.3 | 26 | 89.7 |  |  |  |  |
| Alcohol Habit | Never | 30 | 23.6 | 97 | 76.4 | 0.37 |  |  |  |
|  | Occasional | 20 | 12.2 | 144 | 87.8 |  |  |  |  |
|  | Regul<br>ar | 1 | 20 | 4 | 80 |  |  |  |  |
| Physical | Yes | 23 | 17.4 | 109 | 82.6 | 0.93 | 1.0 | 0.5 | 1.8 |
| Exercise | No | 28 | 17.1 | 136 | 82.39 | 7 | 25 | 59 | 79 |
| Consumption | Yes | 49 | 18.0 | 223 | 82.0 | 0.22 | 2.4 | 0.5 | 10. |
| of fruit habit | No | 2 | 8.3 | 22 | 91.7 | 9 | 17 | 50 | 620 |
| Anyy chronic | Yes | 6 | 8.1 | 68 | 91.9 | 0.01 | 0.3 | 0.1 | 0.8 |
| disease | No | 45 | 20.3 | 177 | 79.7 | 6 | 47 | 42 | 51 |
|  | Yes | 0 | 0 | 7 | 100 | 0.41 | 1.0 | 1.0 | 1.1 |
| Heart Disease | No | 6 | 8.7 | 63 | 91.3 | 6 | 95 | 18 | 78 |
|  | Yes | 0 | 0 | 8 | 100 | 0.38 | 1.0 | 1.0 | 1.1 |
| Arthritis | No | 6 | 8.8 | 62 | 91.2 | 1 | 97 | 19 | 81 |
|  | Yes | 0 | 0 | 9 | 100 | 0.35 | 1.0 | 1.0 | 1.1 |
| Asthama | No | 6 | 9.0 | 61 | 91.0 | 0 | 98 | 19 | 84 |
|  | Yes | 4 | 10.5 | 34 | 89.5 | 0.67 | 2.1 | 0.3 | 12. |
| Hypertension | No | 2 | 5.3 | 36 | 94.7 | 4 | 18 | 64 | 320 |

Above table describes the association of anxiety with several behavioral and health-related factors, displaying the distribution of participants and the p-values, odds ratios (OR) and confidence intervals. Smoking and alcohol use are not significantly associated with anxiety (p > 0.05), however presence of anxiety, was higher in regular smokers (89.7%) and occasional drinkers were 87.8%. Likewise, physical exercise and fruit consumption show no significant effect (p = 0.937 and p = 0.229, respectively) although there is a tendency that anxiety is higher among non-fruit consumers (91.7%). There is a strong association between anxiety and chronic diseases (p=0.016, OR=0.347) thus higher likely hood among the diseased. Certain health conditions such as cardiovascular disease, arthritis, asthma, and hypertension demonstrate higher levels of anxiety; though they are almost all non-significant, indicating that further research is required. The results emphasize chronic disease as an important aspect of anxiety, and longitudinal trends in other variables point towards avenues for further inquiry.

**Table 16.**
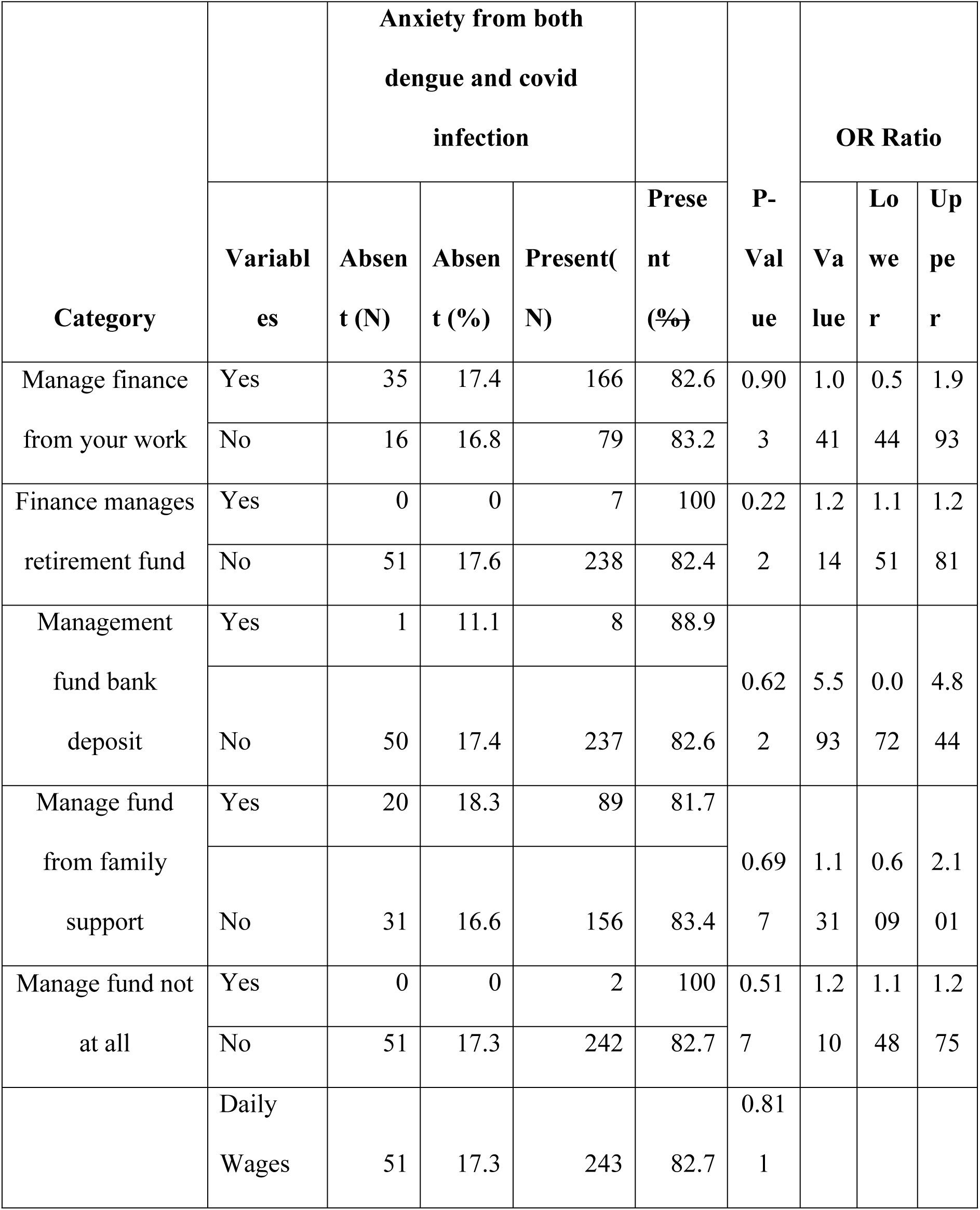
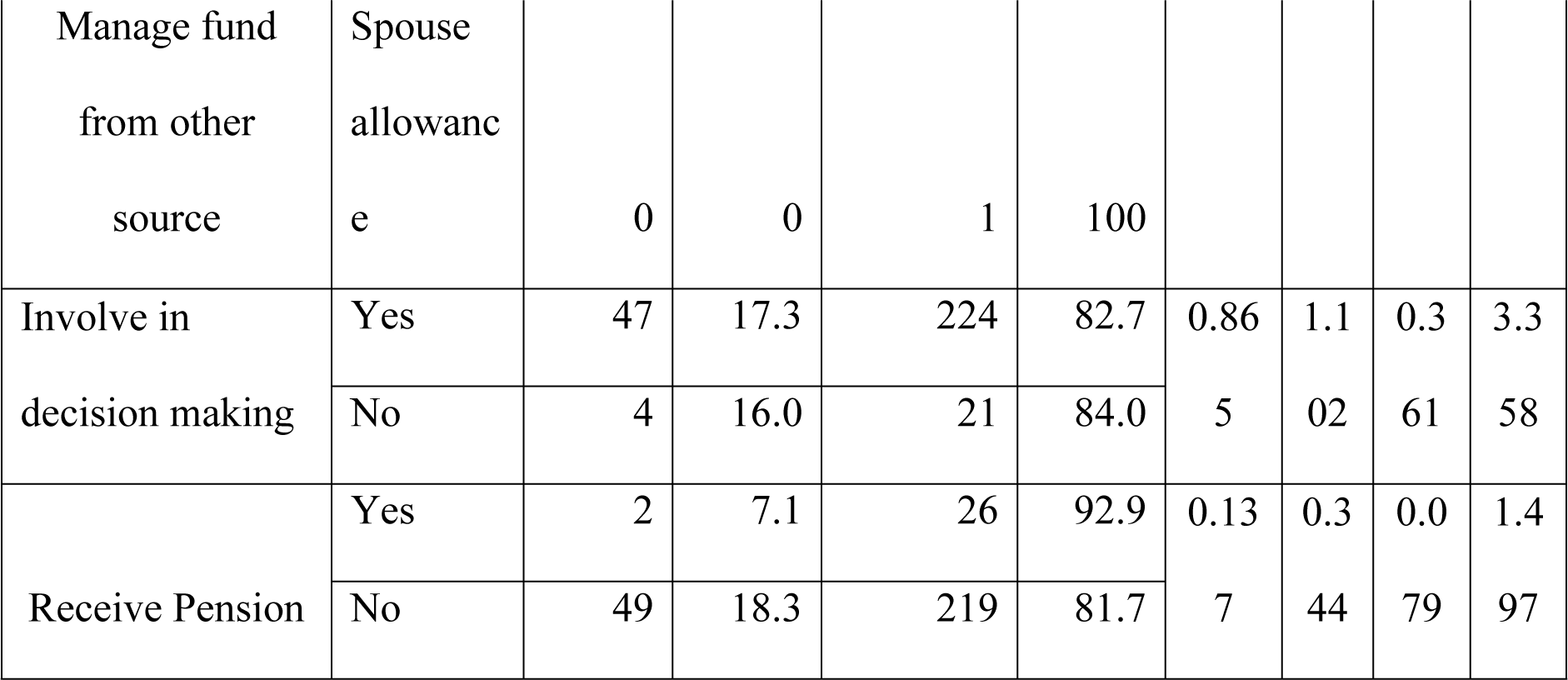
Association between anxiety from both dengue and covid infection with financial choices.

| Category |  | Anxiety from both<br>dengue and covid<br>infection |  |  |  | P-Val<br>ue | OR Ratio |  |  |
| --- | --- | --- | --- | --- | --- | --- | --- | --- | --- |
|  | Variabl<br>es | Absen<br>t (N) | Absen<br>t (%) | Present(<br>N) | Prese<br>nt<br>(%) |  | Va<br>lue | Lo<br>we<br>r | Up<br>pe<br>r |
| Manage finance<br>from your work | Yes | 35 | 17.4 | 166 | 82.6 | 0.90<br>3 | 1.0 | 0.5 | 1.9 |
|  | No | 16 | 16.8 | 79 | 83.2 |  | 41 | 44 | 93 |
| Finance manages<br>retirement fund | Yes | 0 | 0 | 7 | 100 | 0.22<br>2 | 1.2 | 1.1 | 1.2 |
|  | No | 51 | 17.6 | 238 | 82.4 |  | 14 | 51 | 81 |
| Management<br>fund bank<br>deposit | Yes | 1 | 11.1 | 8 | 88.9 | 0.62<br>2 |  |  |  |
|  | No | 50 | 17.4 | 237 | 82.6 |  | 5.5<br>93 | 0.0<br>72 | 4.8<br>44 |
| Manage fund<br>from family<br>support | Yes | 20 | 18.3 | 89 | 81.7 | 0.69<br>7 |  |  |  |
|  | No | 31 | 16.6 | 156 | 83.4 |  | 1.1<br>31 | 0.6<br>09 | 2.1<br>01 |
| Manage fund not<br>at all | Yes | 0 | 0 | 2 | 100 | 0.51<br>7 | 1.2 | 1.1 | 1.2 |
|  | No | 51 | 17.3 | 242 | 82.7 |  | 10 | 48 | 75 |
|  | Daily<br>Wages | 51 | 17.3 | 243 | 82.7 | 0.81<br>1 |  |  |  |
| Manage fund<br>from other<br>source | Spouse<br>allowanc<br>e | 0 | 0 | 1 | 100 |  |  |  |  |
| Involve in<br>decision making | Yes | 47 | 17.3 | 224 | 82.7 | 0.86 | 1.1 | 0.3 | 3.3 |
|  | No | 4 | 16.0 | 21 | 84.0 | 5 | 02 | 61 | 58 |
| Receive Pension | Yes | 2 | 7.1 | 26 | 92.9 | 0.13 | 0.3 | 0.0 | 1.4 |
|  | No | 49 | 18.3 | 219 | 81.7 | 7 | 44 | 79 | 97 |

Above table shows how anxiety connects to different aspects of handling money and making financial choices. It shows how participants are spread out across categories (N, %), along with p-values and odds ratios (OR) that come with confidence intervals. It turns out that handling your finances through your own job or with help from family doesn’t really seem to affect anxiety levels (p = 0.903 and p = 0.697, respectively). On the other hand, relying on a retirement fund or not managing funds at all seems linked to everyone in those groups feeling anxious, but this isn’t statistically significant (p > 0.05). Putting money in the bank or using other sources, like daily wages, doesn’t seem to clearly influence anxiety either (p = 0.622 and p = 0.811, respectively). Being involved in decision-making and getting a pension also don’t show a strong link to anxiety (p = 0.865 and p = 0.137, respectively), even though people who receive a pension tend to be more anxious (92.9%). In general, the data suggests there aren’t strong connections between how people manage their finances and their anxiety levels, although there are some pattern.

**Table 17.** Association between depression from both dengue and covid infection with socio-demographic factors.

| Category | Variables | Depression from both dengue<br>and covid infection |  |  |  | P-Value | OR Ratio |  |  |
| --- | --- | --- | --- | --- | --- | --- | --- | --- | --- |
|  |  | Absent<br>(N) | Absent<br>(%) | Present(N) | Present (%) |  | Value | Lower | Upper |
| Age | 18-40 | 60 | 35.1 | 111 | 64.9 | 0.58 | 1.639 | 0.981 | 2.738 |
|  | 41 and above | 31 | 24.8 | 94 | 75.2 |  |  |  |  |
| Sex | Male | 52 | 34 | 101 | 66 | 0.21 | 1.373 | 0.835 | 2.258 |
|  | Female | 39 | 27.3 | 104 | 72.7 | 1 |  |  |  |
| Religion | Hindu | 70 | 38.3 | 177 | 71.7 | 0.04 | 0.527 | 0.281 | 0.939 |
|  | Non-Hindu | 21 | 42.9 | 28 | 57.1 | 4 |  |  |  |
| Ethnicity | Privileged<br>Janajati | 44 | 23.4 | 144 | 76.6 | 0.001 | 0.397 | 0.238 | 0.659 |
|  | Other Than<br>privileged | 47 | 43.5 | 61 | 56.5 |  |  |  |  |
| Marriage | Married | 54 | 30.7 | 122 | 69.3 | 0.978 | 0.993 | 0.601 | 1.641 |
|  | Other Than<br>Married | 37 | 30.8 | 83 | 69.2 |  |  |  |  |
| Education | Illiterate-Secondary | 27 | 26.5 | 75 | 73.5 | 0.248 | 0.731 | 0.433 | 1.245 |
|  | Above Higher<br>Secondary | 64 | 33 | 130 | 67 |  |  |  |  |
| Household<br>Income | <30000 | 28 | 32.2 | 59 | 67.8 | 0.72 | 0.611 | 0.342 | 1.884 |
|  | >30000 | 63 | 30.1 | 146 | 69.9 | 9 |  |  |  |
| Employment | Yes | 63 | 29.6 | 150 | 70.4 | 0.48 | 0.825 | 0.488 | 1.418 |
|  | No. | 28 | 33.7 | 55 | 66.3 | 6 |  |  |  |
| Household | Yes | 67 | 28.4 | 169 | 71.6 | 0.08 | 0.5 | 0.3 | 1.0 |
| Saving | No | 24 | 40 | 36 | 60 | 2 | 95 | 3 | 72 |

This table shows how depression connects with different background and social factors. We found strong links with religion and ethnicity. People who aren’t Hindu had lower rates of depression (57.1%) than Hindus (71.7%). The odds ratio for this was 0.527 (with a 95% confidence interval between 0.281 and 0.99, and a p-value of 0.044). In the same way, privileged Janajati’s had significantly higher depression rates (76.6%) than other ethnic groups (56.5%). The odds ratio here was 0.397 (95% CI: 0.238–0.659, p = 0.001). Factors like age, gender, marriage status, education, income, job status, and household savings didn’t show a strong link to depression, as their p-values were above 0.05. All in all, these results suggest that a person’s religion and ethnic background play a big role in whether they experience depression in this group of people.

**Table 18.**
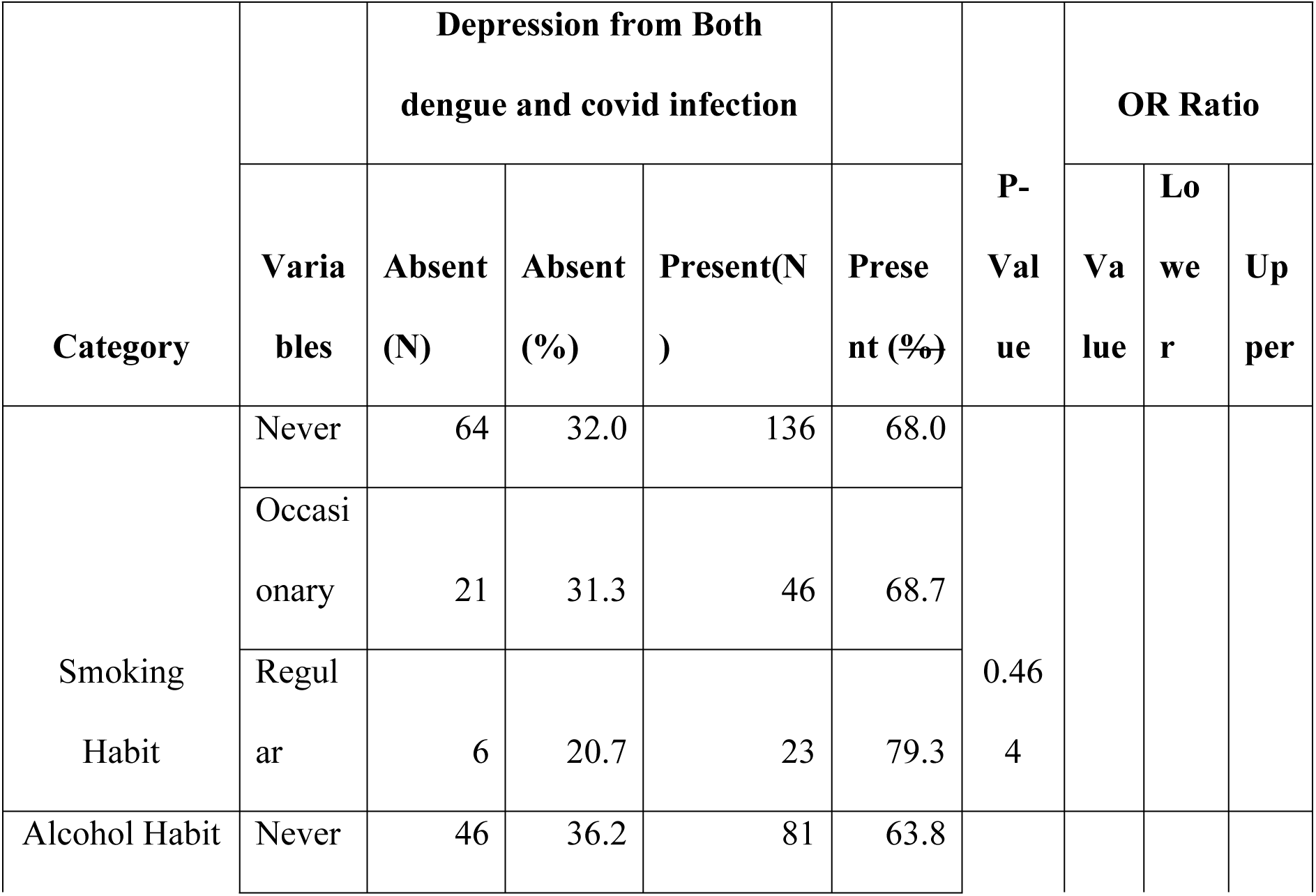
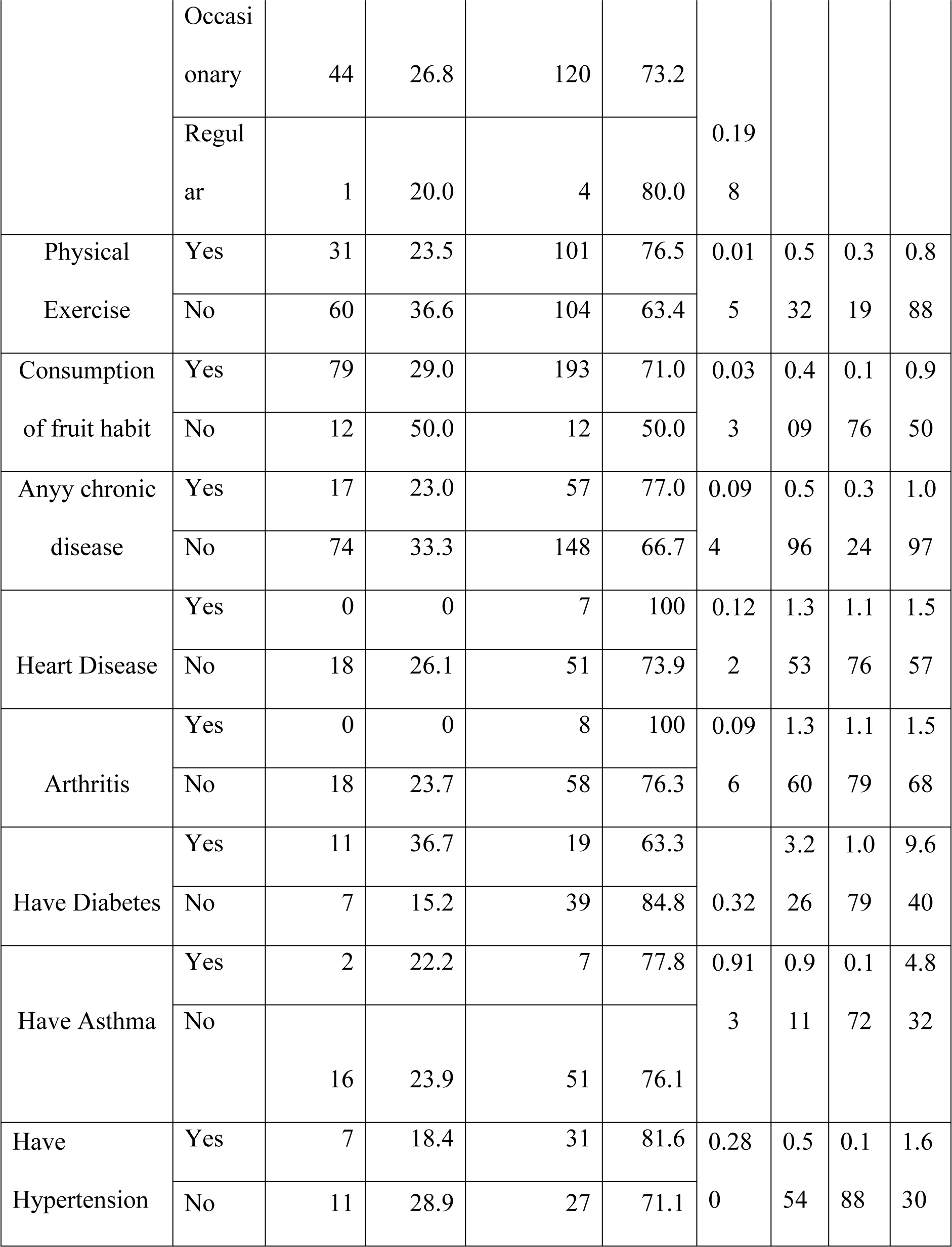
Association between depression from Both dengue and covid infection and health related factors.

Above table shows how depression connects with different behaviors and health factors. It shows the percentage of participants and the results of statistical tests, like p-values and odds ratios (OR) with their confidence intervals. Smoking and drinking habits don’t seem to be strongly linked to depression (p > 0.05), even though more people who smoke regularly (79.3%) and drink regularly (80.0%) tend to have depression. Regular physical activity, on the other hand, is significantly linked to lower depression rates (p = 0.015, OR = 0.532), with those not exercising being more prone to depression. Eating fruit also significantly lowers the chances of depression (p = 0.033, OR = 0.409). While people with chronic diseases like heart disease, arthritis, and hypertension tend to have higher rates of depression, these relationships aren’t statistically significant (p > 0.05). Diabetes exhibits a notable relationship with depression (OR = 3.226), though with limited sample size. The findings suggest significant roles for physical exercise and fruit consumption in mitigating depression, while trends in other variables highlight potential areas for further research.

**Table 19.**
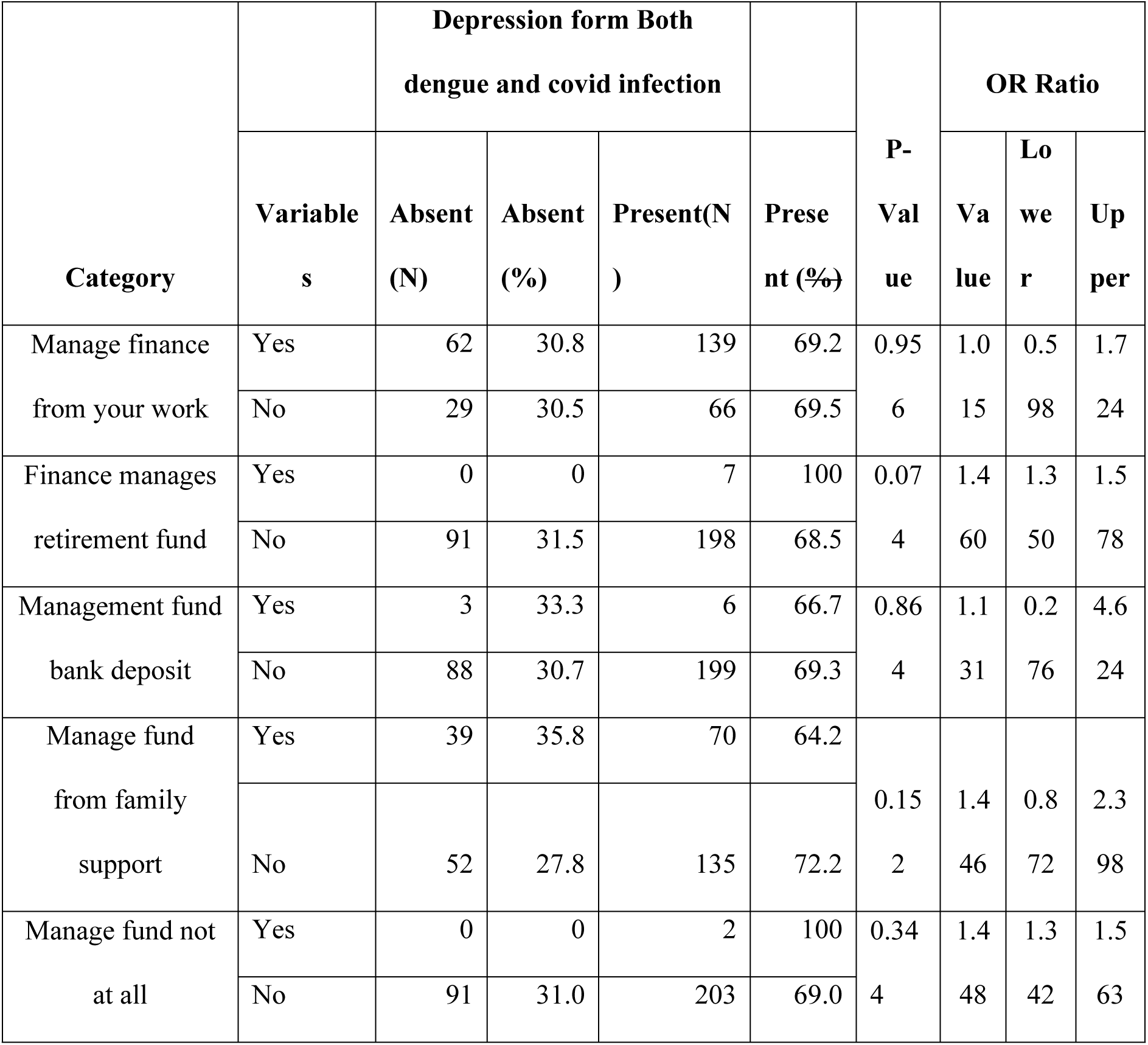
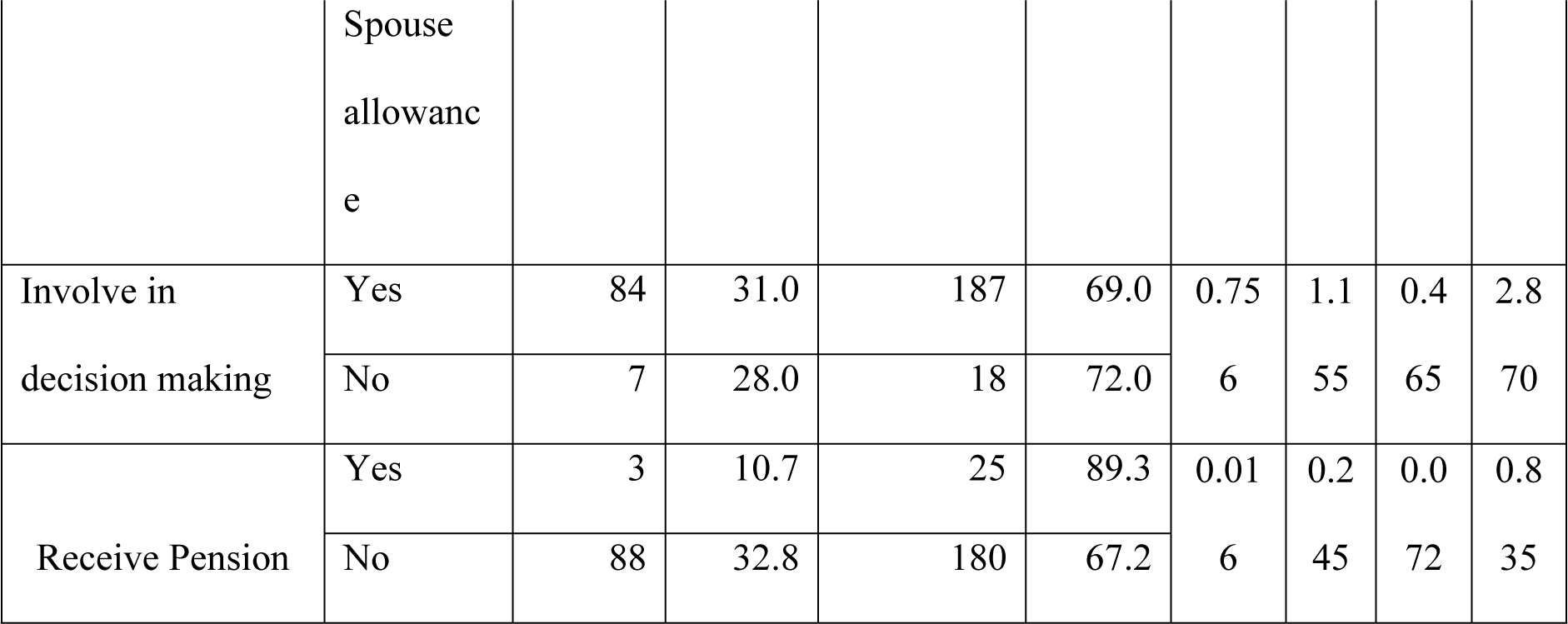
Association between depression from Both dengue and covid infection and financial choices.

This table takes a look at how depression connects to the ways people handle their money. It breaks down the responses from the study participants (using numbers and percentages) and also includes statistical measures like p-values and odds ratios (OR) to show the strength of these connections. The results show that how people manage their money - whether it’s through their own work, retirement savings, or help from family - doesn’t seem to have a strong link to depression (p > 0.05). However, the findings do point out that those who depend only on retirement funds or don’t manage any funds at all experience depression at a rate of 100%. Interestingly, whether or not someone is involved in making decisions about their finances doesn’t really connect to depression either (p = 0.756). But, getting a pension does seem to be tied to a higher chance of being depressed (p = 0.016, OR = 0.245), which might suggest that not having a pension could be somewhat protective against depression. In a nutshell, while receiving a pension does stand out as a factor, the other aspects of managing finances don’t show a strong relationship to depression. This suggests that researchers should probably dig deeper into other economic and psychosocial factors that might contribute to depression.

**Table 20:** Bivariate Analysis to find the association between the level of anxiety due to dengue and dengue with COVID 19.

| Hypothesis | Test Used | Significance Level ($\alpha$) | p-value | Decision | Interpretation |
| --- | --- | --- | --- | --- | --- |
| The distribution of the level of anxiety due to dengue is the same across categories of new anxiety from both groups. | Independent-Samples Mann-Whitney U Test | 0.05 | 0.001 | Null Hypothesis Rejected | There is a significant difference in the distribution of anxiety levels across the groups. |

**Table 21:** Bivariate Analysis to find the association between the level of depression due to dengue and dengue with COVID 19.

| Hypothesis | Test Used | Significance Level ($\alpha$) | p-value | Decision | Interpretation |
| --- | --- | --- | --- | --- | --- |
| The distribution of the level of depression due to dengue is the same across categories of new depression from both groups. | Independent-Samples Mann-Whitney U Test | 0.05 | 0.000 | Null Hypothesis Rejected | There is a significant difference in the distribution of depression levels across the groups. |

$\alpha= 0.05$ , CI= 95%

As the data followed non normal distribution, Mann Whitney U test is performed between level of anxiety due to dengue and level of anxiety due to infection of both dengue and COVID 19. The finding from the result showed the significant association between dependent and independent variables (p=0.001) which is less than the significant level α <0.05 suggesting that dengue related anxiety varies meaningfully across various groups.

When performing Mann-Whitney U Test, the result revealed significant difference at a statistically significant difference (p = .001), which is below the significance level of α = 0.05. Suggesting level of depression due to dengue significantly varies across different groups.

### Multivariate Analysis

Multivariate analysis was carried to show true association between dependent and independent variables. Those variables that were significant in bivariate analysis were taken into account for multivariate analysis. Multivariate Analysis (binomial regression) between independent variables and level of depression due to both dengue and covid 19 infections.

**Table 22.** Association between dependent and independent variables.

| Category | P-value | AOR | 95% C.I. |  |
| --- | --- | --- | --- | --- |
|  |  |  | Lower | Upper |
| Completed age | .205 | 1.011 | .994 | 1.028 |
| Ethnicity | .573 | 1.114 | .765 | 1.624 |
| Level of Education | .479 | .846 | .533 | 1.344 |
| Have habit of smoking | .392 | .392 | .046 | 3.346 |
| Have habit of alcoholism | .281 | 1.332 | .791 | 2.244 |

Cox & Snell R Square = 0.205, Nagelkerke R Square= $R^2 = 0.308$

The value of Nagelkerke R² of the above table suggests the model accounts for about 30.8% of the variation in depression levels. The p-value in the table shows no significant relationship of independent variables like age, ethnicity, level of education, habit of alcoholism with depression is found.

**Table 23.** Multivariate Analysis between independent variables and level of Anxiety due to both dengue and covid 19 infections.

| Category | P-value | AOR | 95% C.I.for EXP(B) |  |
| --- | --- | --- | --- | --- |
|  |  |  | Lower | Upper |
| Age | .255 | .607 | .256 | 1.436 |
| Ethnicity | .004 | 2.583 | 1.364 | 4.890 |
| Education | .067 | 2.290 | .943 | 5.564 |
| Have habit of Smoking | .049 | 0 | 0 |  |
| Have habit of alcohol consumption | .489 | 2.365 | .207 | 27.036 |
| Have any chronic disease | .384 | 1.591 | .559 | 4.526 |

Cox & Snell R Square = 0.093 , Nagelkerke R Square= $R^2 = 0.154$

This model shows **Nagelkerke R² = 0.154**. The model explains about 15.4% of the variance in anxiety levels. Smoking habit and ethnicity show significant association with presence of depression. Age, habit of alcohol and have any chronic disease does not significantly impact anxiety level.

## DISCUSSION

This research study aims to assess the anxiety and depressive symptoms in patients from the Kathmandu Valley with post-dengue and dengue plus COVID-19 simultaneously. Additionally, it aimed to analyze the factors contributing to these mental health issues including demographic, socioeconomic, and health related aspects. The results shed further light on the relationship between infectious diseases and mental health, showcasing the need for multidisciplinary healthcare services.

There were notable relationships between anxiety and certain demographic characteristics such as age, ethnicity, and education level. Patients aged 41 years and older were more anxious (90.4%) compared to younger patients (77.2%), resulting in an odds ratio (OR) of 2.782 (p = 0.003). Ethnic differences were noted whereby privileged Janajatis were relatively less anxious (88.3%) than others (73.7%), with an OR of 0.361 (p = 0.001). Additionally, having a higher education was said to have an impact on anxiety levels, with lower degree qualifications increasing the odds of anxiety (OR 0.35, p 0.005). These results corroborate pre-existing literature, which emphasizes the impact of education and age on mental health during the pandemic (17).

The heightened anxiety and depression levels among the respondents with the co-infected group in our study align with prior studies on post-viral psychological sequelae. An epidemiological study conducted in Bangladesh showed that individuals with dengue and COVID-19 co-infection were at significantly higher risk of having severe health conditions like brain fog, mood disturbances, and fatigue. (18)

A case-control study conducted in UK found no significant differences in the level of anxiety and depression between COVID-19 patients with and without neuropsychiatric complications, but the report supported that increased psychological burden in co-infected cases. (19)

A study conducted in Romania demonstrated higher postnatal anxiety among COVID-19- positive women compared to uninfected controls. The study indicated that the infection of COVID-19 played a significant role in causing anxiety in vulnerable populations. (20)

A prospective cohort study showed that 44.4% of cases and 17.1% of controls showed that stress score increased at the 3^rd^ month (RR 1.87, 95% CI [1.01-3.4]. (**21**)

Nearly 60% of the patients had anxiety, and 62.2% of the patients had depression due to infection of dengue. (22)

A significant increase in depression across all timeframes was observed (<3 months [aSHR 1.90, 95% CI 1.20–2.99], 3–12 months [aSHR 1.68, 95% CI 1.32–2.14], and >12 months [aSHR 1.14, 95% CI 1.03–1.25]). Sleep disorder risk was higher only during 3–12 months (aSHR 1.55, 95% CI 1.18–2.04). Increased risk of anxiety disorders in 3 months (aSHR 2.14, 95% CI 1.19–3.85) and persistent risk of depression across all periods was found. (23)

The variables like age over 40, lower educational level, and marginalized ethnicity were found significantly associated with higher anxiety and depression, The finding is similar to the study indicating older adults and socially disadvantaged groups are more susceptible to pandemic-related mental distress. (19,20)

In our study, health related behavior such as insufficient fruit intake and lack of physical activity were found significantly associated with depression. These modifiable variables may influence through inflammatory or neurobiological pathways, as poor diet and lack of physical activity is significantly related with altered cortisol and cytokine profile-contributors of affective disorders. The study revealed that history of chronic illness was a significant predictor of anxiety suggesting comorbid conditions can exacerbate mental responses to infectious disease. Although in our study the conditions like heart disease, arthritis, and asthma did not show a significant association, the heightened anxiety among these groups suggests for more powerful group analysis. (18)

Those receiving a pension appeared protective against depression, suggesting financial security acts as a buffer against pandemic-induced mental health problems. Conversely, lack of saving and financial uncertainty indicate elevated psychological distress, though statistical significance was not met during multivariate analysis. (20)

The analysis of depression revealed significant links with religion and ethnicity. Non-Hindu participants exhibited lower depression prevalence (57.1%) than Hindus (71.7%), with an OR of 0.527 (p = 0.044). Privileged Janajatis were more likely to experience depression (76.6%) compared to other ethnicities (56.5%) with an OR of 0.397 (p = 0.001). Behavioral factors such as physical exercise and fruit consumption significantly mitigated depression (p = 0.015 and p = 0.033, respectively), corroborating studies on the mental health benefits of healthy lifestyles (24).

However, multivariate analyses suggested limited explanatory power, with Nagelkerke R² values of 15.4% for anxiety and 30.8% for depression. Smoking and ethnicity were the only significant predictors in the anxiety model, while other factors like age and chronic diseases were not significant. These findings suggest the complex and multifactorial nature of anxiety and depression, influenced by a combination of individual, social, and contextual factors.

The findings underscore the mental health burden associated with infectious diseases like dengue and COVID-19. Integrated mental health interventions addressing demographic and behavioral risk factors are essential for mitigating this burden. Future research should explore longitudinal impacts and potential protective mechanisms to enhance resilience in affected populations.

### Conclusion

This study demonstrates that prior infections of dengue, with and without COVID-19, significantly influence mental health outcomes in adults in the Kathmandu Valley. Anxiety and depression levels were markedly higher in specific demographic and socioeconomic groups, including older adults, certain ethnicities, and individuals with lower educational attainment. Behavioral factors such as smoking, physical activity, and diet further shaped mental health outcomes, with healthy behaviors providing protective benefits. These findings highlight the necessity of addressing mental health as part of infectious disease management.

## Data Availability

The cleaned data entered in SPSS will be provided upon request. We also can provide you the evidence of data collection (hard copies too)

## Acknowledgements

We are thankful to the Asian College for Advanced Studies for granting me the opportunity to conduct this research. Your support and encouragement have been invaluable throughout this process.

Special thanks go to Anita Ghimire, Shanti Ghimire, Srijana Shrestha, Rubi Shrestha, and Asmita Sanjel, along with the dedicated Public Health graduates, for their tireless efforts in collecting data from Lalitpur, Bhaktapur, and Kathmandu. Last but not the least the support of Bikesh Deshar, our sincere student, also supported in the finalization of the report; we are very thanking you towards him. Your hard work and commitment have significantly contributed to the success of this research.

Thank you all for your contributions and support.

---

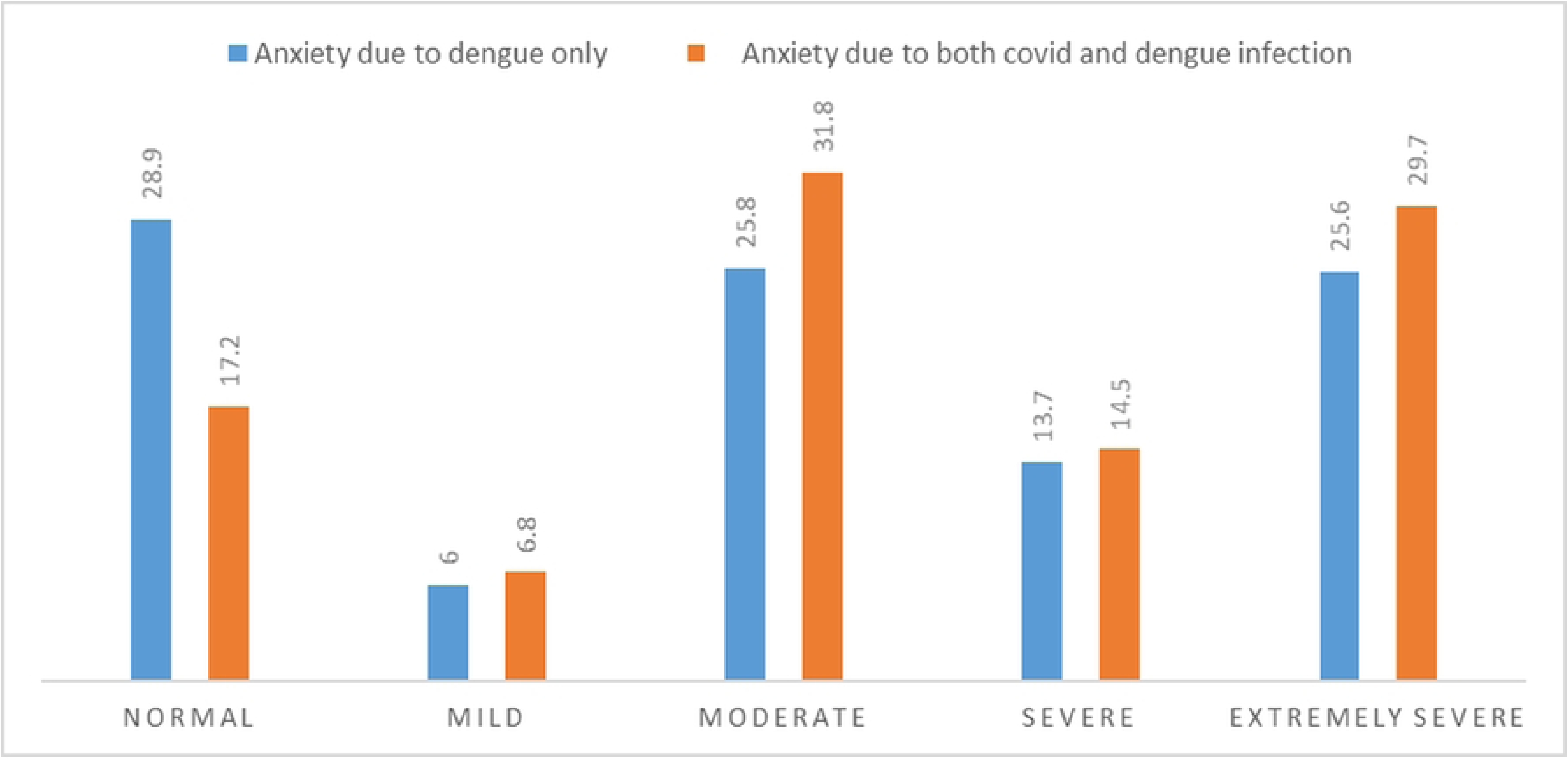

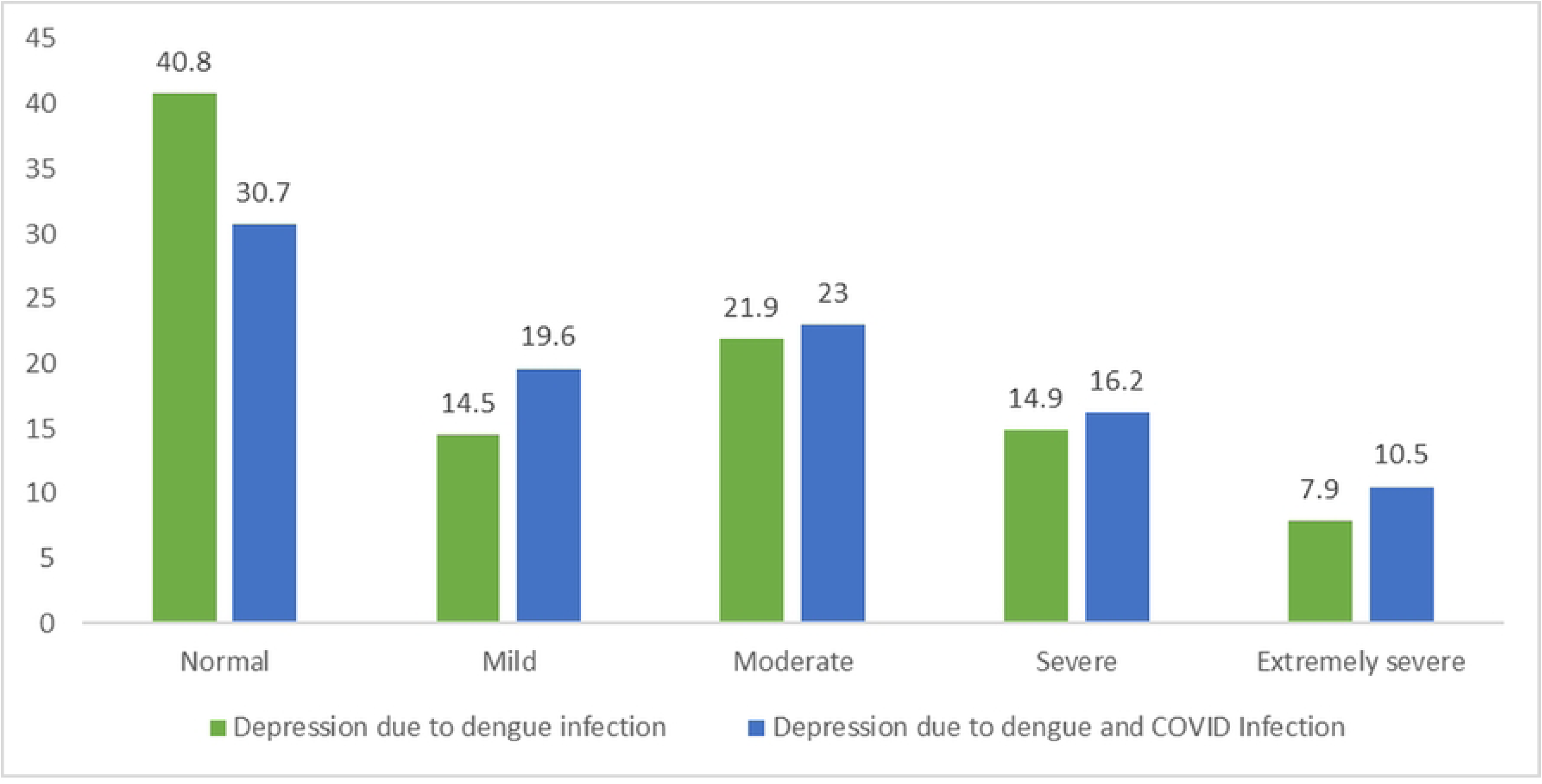

